# Multidimensional factors predicting exclusive breastfeeding in Ethiopia: evidence from a meta-analysis of studies in the past 10 years

**DOI:** 10.1101/19002857

**Authors:** Tesfa Dejenie Habtewold, Aklilu Endalamaw, Shimels Hussien Mohammed, Henok Mulugeta, Getenet Dessie, Getachewu Mullu Kassa, Yared Asmare, Mesfin Tadesse, Yihun Mulugeta Alemu, Niguse Tadesse Sharew, Abera Kenay Tura, Balewgizie Sileshi Tegegne, Sisay Mulugeta Alemu

## Abstract

**Background:** In Ethiopia, the prevalence of exclusive breastfeeding (EBF) is 60.1%, which is lower than the national Health Sector Transformation Plan 2016-2020, National Nutrition Program 2016–2020 and WHO global target. This may be attributed to multidimensional factors.

**Objective:** The aim of this meta-analysis was to investigate the association between EBF and educational status, household income, marital status, media exposure, and parity in Ethiopia.

**Methods:** Databases used were PubMed, EMBASE, Web of Science, SCOPUS, CINAHL and WHO Global health library, and key terms were searched using interactive searching syntax. It was also supplemented by manual searching. Observational studies published between September 2000 and March 2019 were included. The methodological quality of studies was examined using the Newcastle-Ottawa Scale (NOS) for cross-sectional studies. Data were extracted using the Joanna Briggs Institute (JBI) data extraction tool. To obtain the pooled odds ratio (OR), extracted data were fitted in a random-effects meta-analysis model. Statistical heterogeneity was quantified using Cochran’s Q test, τ^2^, and I^2^ statistics. Additional analysis conducted includes Jackknife sensitivity analysis, cumulative meta-analysis, and meta-regression analysis.

**Results:** Out of 553 studies retrieved, 31 studies fulfilled our inclusion criteria. Almost all studies were conducted on mothers with newborn less than 23 months. Maternal educational status (OR = 1.39; p = 0.03; 95% CI = 1.03 - 1.89; I^2^ = 86.11%), household income (OR = 1.27; p = 0.02; 95% CI = 1.05 - 1.55; I^2^ = 60.9%) and marital status (OR = 1.39; p = 0.02; 95% CI = 1.05 - 1.83; I^2^ = 76.96%) were found to be significantly associated with EBF. We also observed an inverse dose-response relationship of EBF with educational status and income. Significant association was not observed between EBF and parity, media exposure and paternal educational status.

**Conclusions:** In this meta-analysis, we depicted the relevant effect of maternal education, income, and marital status on EBF. Therefore, multifaceted, effective, and evidence-based efforts are needed to increase national breastfeeding rates in Ethiopia.

## Background

Optimal breastfeeding is an essential element for the satisfactory growth and development of infants and children by reducing and/or preventing morbidity and mortality related to malnutrition and infection.^1-4^ Exclusive breastfeeding (EBF) significantly lowers the risk of sepsis, diarrhea and respiratory infections as evidenced by systematic review and meta-analysis.^5^ Recent estimates suggest that optimal breastfeeding could prevent around 12% deaths in under-5 children every year, representing around 800,000 lives in low- and middle-income countries. ^6^ Breastfeeding has also economic and environmental advantages. ^7^ EBF prevents 20% to 22% of neonatal deaths.^2,3,8^ Consequently, the Innocenti Declaration, UNICEF, World Health Organization (WHO), British Paediatric Association, American Dietetic Association, and American Academy of Pediatrics have recommended EBF for the first six months.^9^ Despite these facts, every new-born have the right to exclusively breastfeed for the first six months^10,11^ but it is suboptimal in developed and developing countries.^12,13^ Non-exclusive breastfeeding in the first 6 months of life resulted in 1.4 million deaths and 10% of the disease burden in children younger than 5 years of age.^14^ Overall, in low- and middle-income countries, only 37% of children younger than 6 months of age are exclusively breastfeed.^15^ Based on secondary data analysis of 32 Demographic and Health Surveys conducted in Sub-Saharan African countries since 2010, the prevalence of EBF is 41.0%.^16^ In Ethiopia, breastfeeding is a universal practice with 96% of children breastfed at some point in time. ^17^ A meta-analysis study^18^ from our research group shows that the prevalence of EBF is 60.1%, which is higher than the Global Nutrition Target 5 of 50 % EBF coverage by 2025.^91^

Exclusively breastfeeding has been associated with parental educational status, marital status, household income, number of children, maternal age, child age, antenatal care, place of delivery, postnatal care, gender, birth order, employment status, mode of delivery and timely initiation of breastfeeding.^8,12,19-23^ However, a prospective study shows that an absence of significant association between EBF and parental education, living conditions, antenatal care follow-up, birth weight, culture, postnatal breastfeeding advice, previous breastfeeding exposure and mothers’ employment.^24^ Likewise, a systematic review of 48 studies from 14 low- and middle-income US Agency for International Development (USAID) Ending Preventable Child And Maternal Deaths (EPCMD) priority countries identified 16 barriers of EBF.^25^ Based on a pooled Demographic and Health Survey data sets collected in nine Sub Saharan African countries, maternal educational status, maternal employment, ANC, PNC and place of delivery were significantly associated with EBF.^26^

In Ethiopia, a systematic review of 32 studies identified 26 factors that significantly associated with EBF and categorized into four groups – proximal, proximal-intermediate, distal-intermediate and distal. Proximal factor includes maternal educational status, mother’s knowledge on EBF, guidance and counseling, access to health facility, poor attitude, and age of newborn. Proximal-intermediate factor includes place of delivery, timely initiation of breastfeeding, intention or plan to breastfeeding, mode of delivery, breast complication, breastfeeding self-efficacy and outcome expectancy, and sex of newborn. Distal-intermediate factor includes ANC, PNC, colostrum feeding, and prelacteal feeding. Finally, distal factor includes paternal educational status, household income, marital status, parity, husband support, maternal age, place of residence and family size.^18^ The follow-up meta-analyses confirmed that maternal occupational status, gender of newborn, age of newborn, guidance and counseling on breastfeeding, colostrum discarding, ANC, PNC, vaginal delivery, health institution delivery, timely initiation of breastfeeding significantly increased the odds of EBF.^18,27,28^

There is no doubt regarding the multiple benefits of breastfeeding for infants and society, in general. As a result, global initiatives such as the International Code of Marketing of Breast-milk Substitutes (referred to hereafter as ‘the Code’), the Innocenti Declaration, the Baby-Friendly Hospital Initiative (BFHI), the Millennium Development Goals, and the Global Nutrition Targets 2025 and Sustainable Development Goals have developed.^26,29^ The Ethiopian government has also endorsed and implemented these programs to reduce infant and child mortality and morbidity. A national nutrition strategy and program (NNP) has been developed and implemented in a multi-sectoral approach. The Health Sector Development Program (HSDP IV) has integrated nutrition into the Health Extension Program to improve the nutritional status of mothers and children through the Community Based Nutrition program (CBN), Health Facility Nutrition Services, and Micronutrient Interventions and Essential Nutrition Actions / Integrated Infant and Young Feeding Counseling Services.^30^ Despite the reduced under-five mortality, the rate of malnutrition, rate of stunting and rate of underweight^31^, the rate of EBF has fallen short of the Health Sector Transformation Plan 2016-2020^30^, National Nutrition Program 2016–2020^32^ and WHO global target. These drawbacks call for the need to assess multidimensional factors that affect EBF. Thus, the aim of this meta-analysis was to investigate the association between parental educational status, household income, parity, exposure to media and marital status with EBF in Ethiopia.

## Methods

### Protocol and registration

This systematic review and meta-analysis was conducted based on the registered (CRD42017056768) and published protocol.^33^ Based on the authors’ decision, the following changes were made to the published protocol.^33^ Joanna Briggs Institute (JBI) tool^34^ was used to extract the relevant data. Furthermore, cumulative meta-analysis and mixed-effects meta-regression analysis were done to reveal the trends of evidence and identify possible sources of between-study heterogeneity respectively.

### Measurement variables

The outcome measurement was exclusive breastfeeding (EBF), which is the proportion of children born in the last two years who were breastfeed only breastfeed for the first six months after delivery.^35^ Maternal educational status, paternal educational status, household income, marital status, media exposure, and parity were selected predicting factors. The selection of these factors was guided by our previous systematic review findings.^18^ Educational status represents the highest schooling grade achieved as per the Ethiopian educational system and categorized as ‘uneducated’ (including those who able to read and write without formal schooling), ‘primary’ (grades 1 to 8) and ‘secondary and above’ (grades 9 or above). Household income was categorized as ‘high’, ‘medium’ and ‘low’. Because of substantial inconsistency in the reported household income, we used a harmonized qualitative way of classification of income for all included studies that reported at least three categories of household income or wealth index based on authors educational judgment. Marital status was categorized as ‘currently married’ and ‘others’ (i.e. single, divorced, widowed). Media exposure represents exposure to or ownership of any print media (newspaper, leaflet, brochure) and broadcasting (radio and television) and categorized as ‘yes’ and ‘no’. This does not include Facebook, email, YouTube, or WhatsApp as most mothers do not have access to internet. If studies reported accessibility or exposure to more than one media tools, we extracted data on radio (as ‘yes’) followed by television and print media. Parity refers to the total number of births after 28 weeks and was categorized as ‘primipara’ if the mothers have only one birth and ‘multipara’ if the mothers have at least two births.

### Search for literature

Databases searched were PubMed, EMBASE, Web of Science, SCOPUS, CINAHL and WHO Global health library. The interactive searching syntax was developed for all databases in consultation with librarian, experts on literature searching (Supplementary file 1). We further manually searched the table of contents of Ethiopian Journal of Health Development, Ethiopian Journal of Health Sciences, Ethiopian Journal of Reproductive Health, International Breastfeeding Journal, BMC Pregnancy and Childbirth, BMC Public Health, BMC Paediatrics, Nutrition Journal and Italian Journal of Paediatrics. We also searched cross-references and grey literature on Addis Ababa University institutional research collection repository database. The last search was done in March 2019.

### Inclusion and exclusion criteria

The studies were included when they met the following inclusion criteria: (1) observational studies, such as cross-sectional, case-control and cohort studies; (2) studies conducted in Ethiopia; (3) studies reported on the association between EBF (i.e. operationalized based on the WHO definition) and maternal and paternal educational status, household income (at least three categories must be reported), marital status, media exposure (not exposed or no access to media category must be reported) and parity; (4) studies published from September 2000 (i.e. the time when last revision of the global breastfeeding recommendations occurred) to March 2019. Program evaluation reports, systematic reviews and meta-analyses, qualitative studies and studies on mothers with medical conditions including HIV/AIDS and pre-term or ill health newborn were excluded.

### Screening and selection of studies

Initially, all identified studies were exported into RefWorks citation manager version 4.6 for Windows. Afterward, duplicate studies were deleted from further screening. Next, a pair of reviewers independently screened the abstracts and titles using Microsoft Excel spreadsheet for relevance, their compliance with our measurements of interest and against our inclusion criteria. Based on Cohen’s Kappa inter-rater reliability test, the agreement between the two reviewers was 0.76 indicating substantial agreement between the two reviewers. Disagreements on the inclusion of titles or abstracts were solved through discussion and consensus. After removing irrelevant studies, full text of selected abstracts were downloaded and reviewed for further eligibility. PRISMA flow diagram was also used to illustrate the screening and selection processes of studies.^36^ Finally, two independent reviewers (TD and SM) extracted the following information from each included studies using Joanna Briggs Institute (JBI) data extraction tool^34^: author name, publication year, residence, study design, study population, number of participants, source of funding, and observed data. If funding source was not explicitly mentioned, we reported as ‘not mentioned’, whereas ‘no funding’ category was used if the author explicitly mentioned there is no funding or funding not applicable. If funding was not given directly but through other donors, the original donor was mentioned. Newcastle-Ottawa Scale (NOS) for cross-sectional studies was used to examine the quality of studies and the potential risk of bias.^37^ The scoring system, selection of cut-off value and interpretation used in this meta-analysis were published in our protocol.^33^ Furthermore, we reported the results in compliance with the recommendation of Preferred Reporting Items for Systematic review and Meta-analysis (PRISMA) statement (Supplementary file 2).^36^

### Statistical analysis

To obtain the pooled odds ratio (OR), extracted data were fitted in a random-effects meta-analysis model. In addition, a cumulative meta-analysis was done to illustrate the trend of evidence regarding the effect of predictors on EBF and interpreted as stable, steadily increased/decreased, slightly increased/decreased or dramatically increased/decreased. Publication bias was assessed by subjective evaluation of the funnel plot, and then, we performed Egger’s regression statistical test to objectively confirm the presence of significant publication bias at p-value ≤ 0.01.^38^ We used Cochran’s Q test to test heterogeneity, τ^2^ to estimate the amount of total/residual between-study variance, and I^2^ statistics to measure the proportion of total variation between study due to heterogeneity.^39^ Clinical and methodology heterogeneity were also carefully evaluated. Factors attributed to between-study heterogeneity were investigated using mixed-effects meta-regression analysis using region, residence, sample size and publication year as covariates.^40^ The residual amount of heterogeneity was subtracted from the proportion of heterogeneity and divided by the total amount of heterogeneity to obtain the total amount of heterogeneity (R^2^) explained by covariates. Omnibus test of moderators was applied to assess the moderation effect of these covariates. Meta-regression analysis was done only when heterogeneity threshold (I^2^) was ≥ 80%. Jackknife sensitivity analysis was done to examine the influence of outlier studies on the pooled OR estimate, a significance level of estimate and between-study heterogeneity.^41^ The study was excluded when the pooled OR estimate increased or decreased by one and changes the significance level after lifting out that study from the meta-analysis. Because of the small number of studies available for some variables, the change in heterogeneity threshold was not considered as a primary criterion to detect and excluded the outlier study. The data were analyzed using “metafor” packages in R software version 3.2.1 for Windows.^40^

## Results

### Search results

Of 553 studies retrieved through electronic databases and manual searching, full-text of 41 studies were reviewed. 10 studies were excluded after the full-text review: three studies reported only the prevalence of EBF, and the other seven studies did not report the selected factors of our interest. Detailed characteristics of studies that reported each variable was presented in Table 1. All studies had good methodological quality (NOS score ≥7). One study reported more than one influencing factors. The screening and selection process has been illustrated below using the PRISMA flow diagram (Figure 1).

**Table 1:**
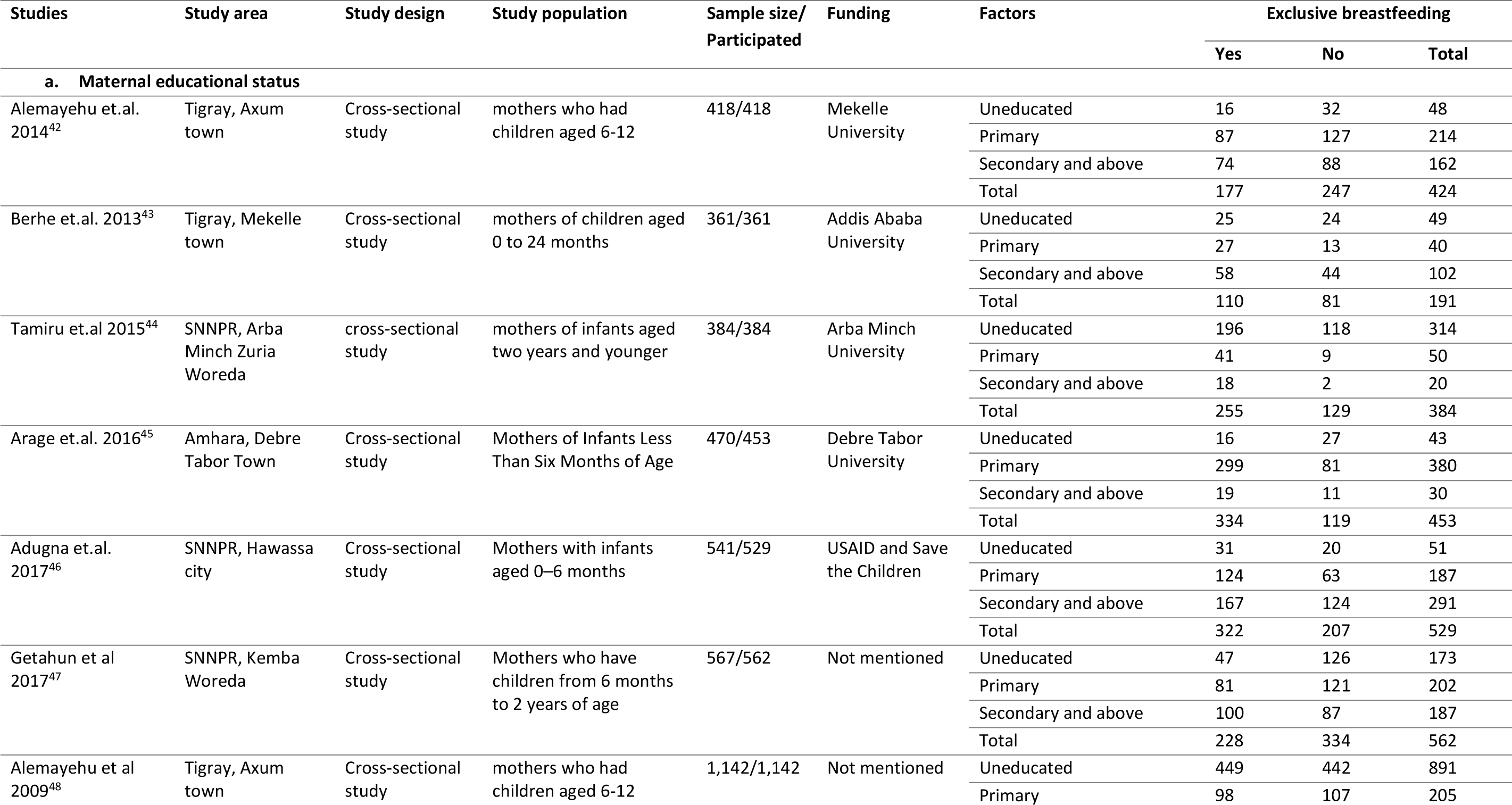
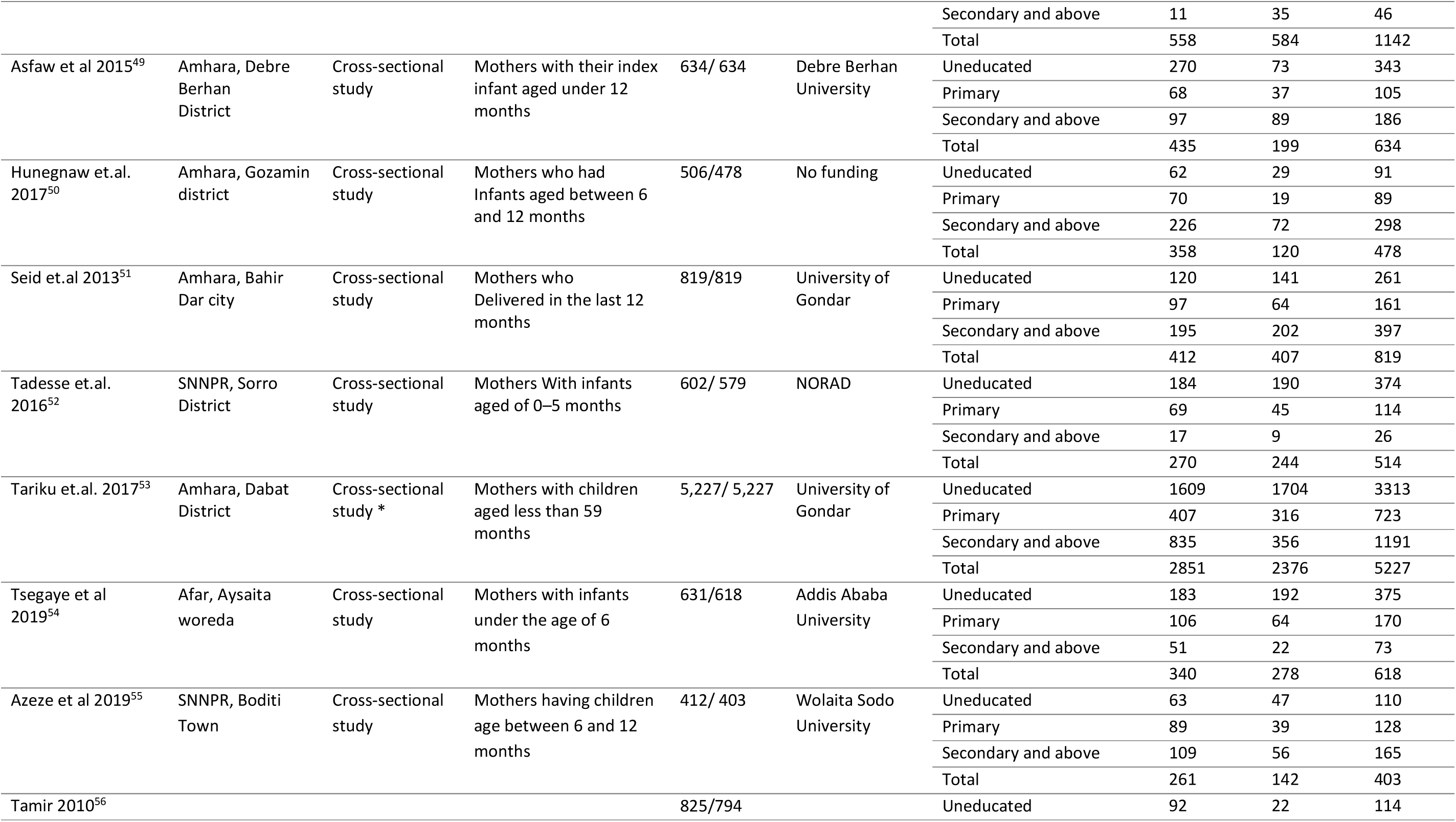
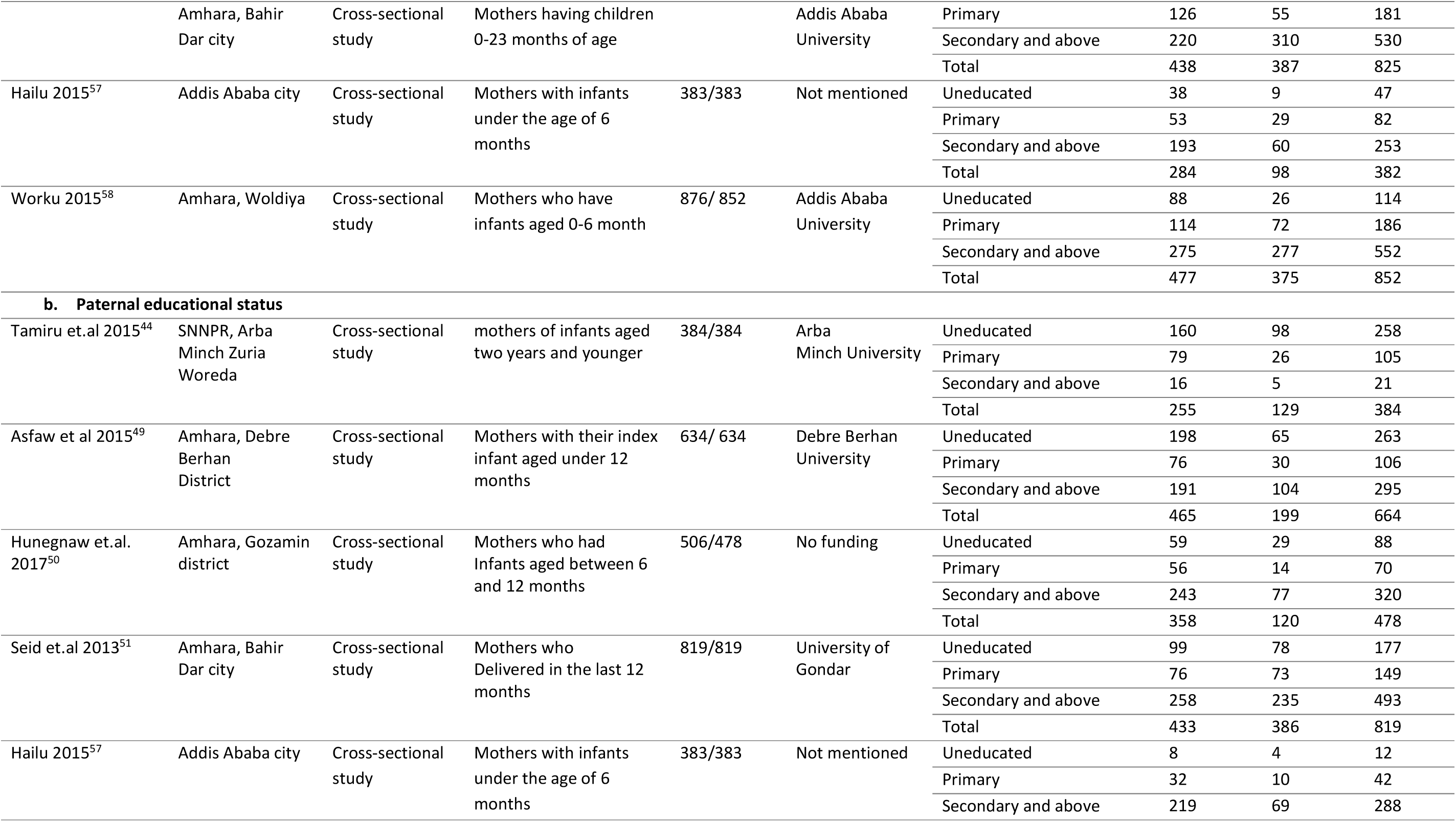
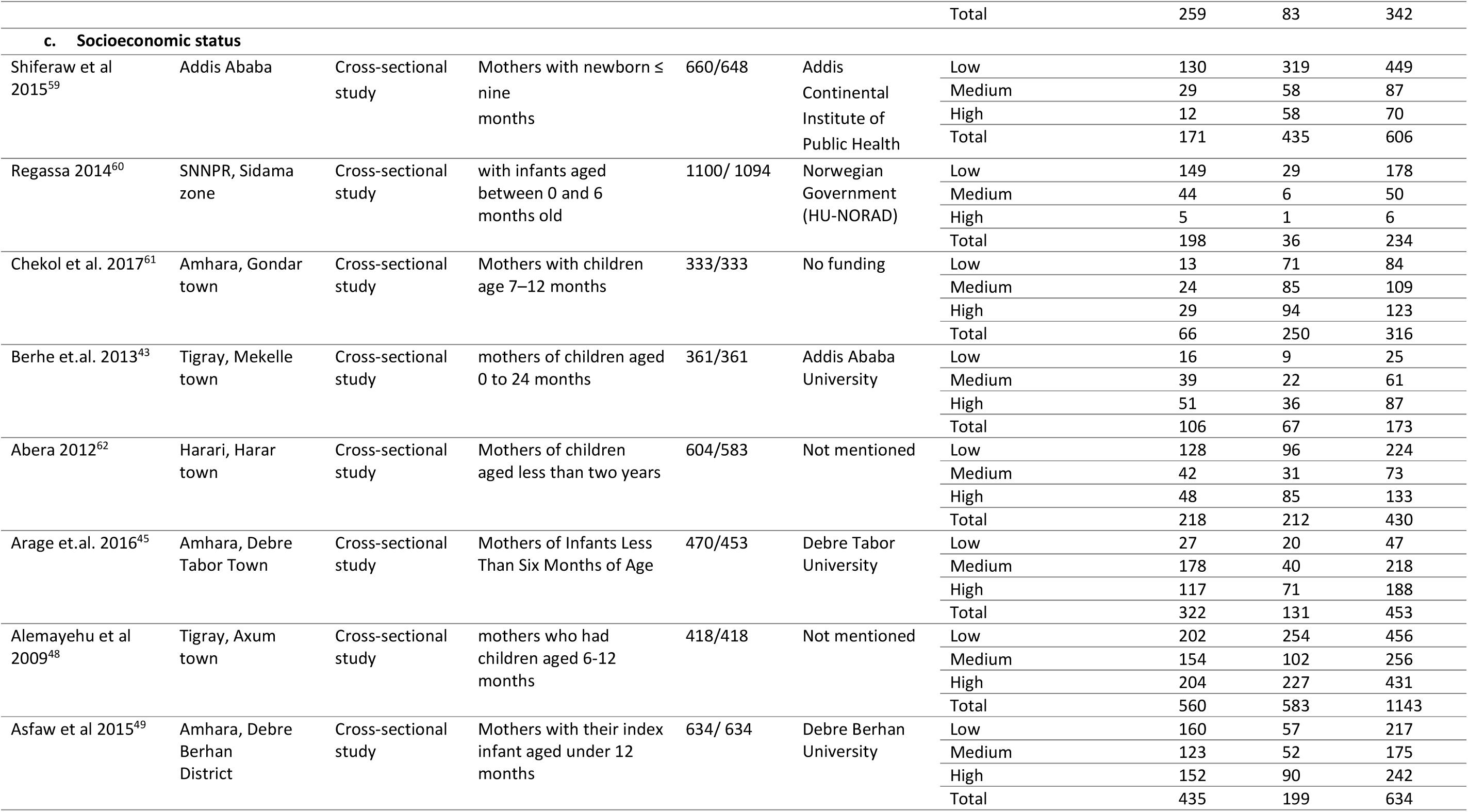
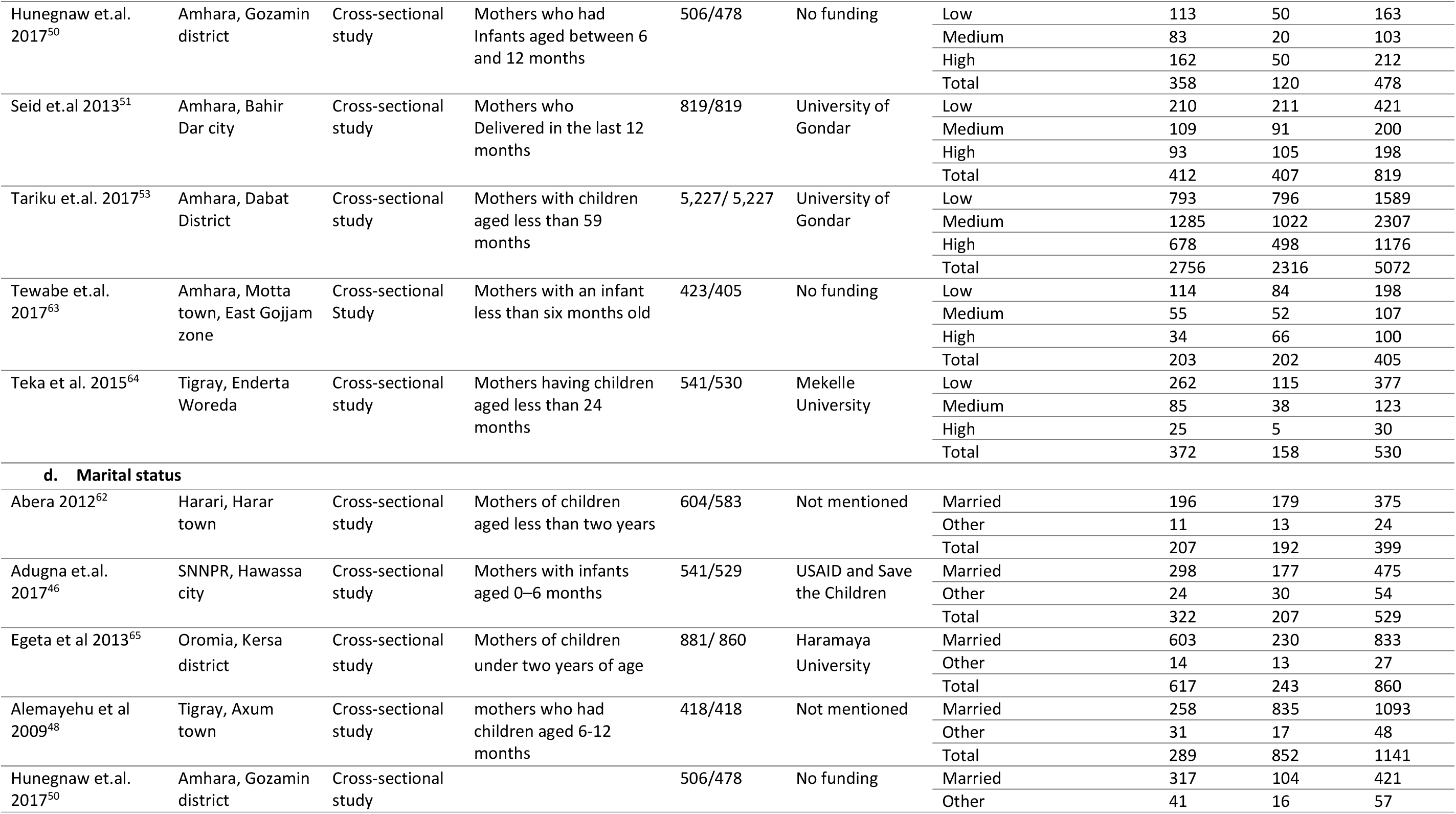
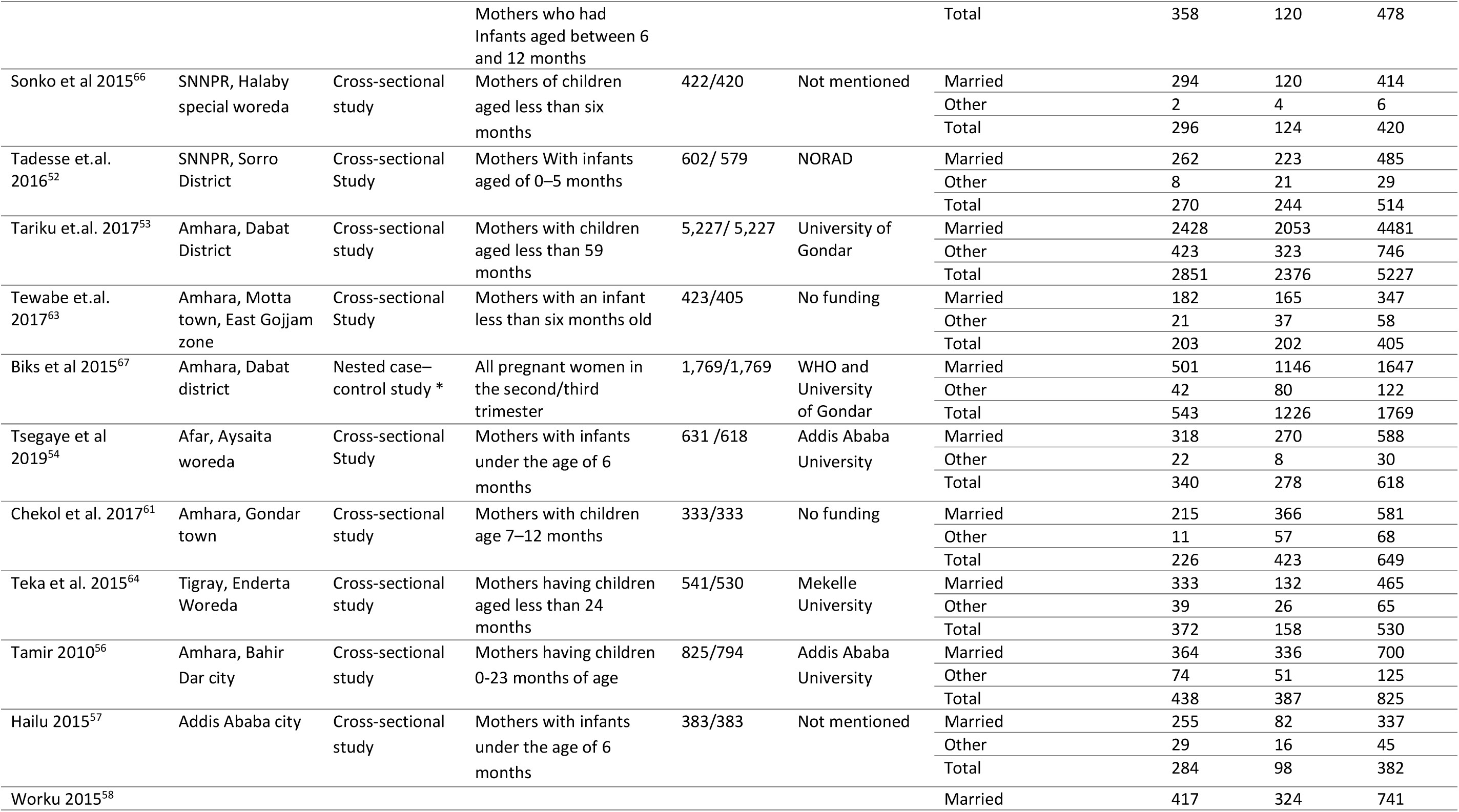
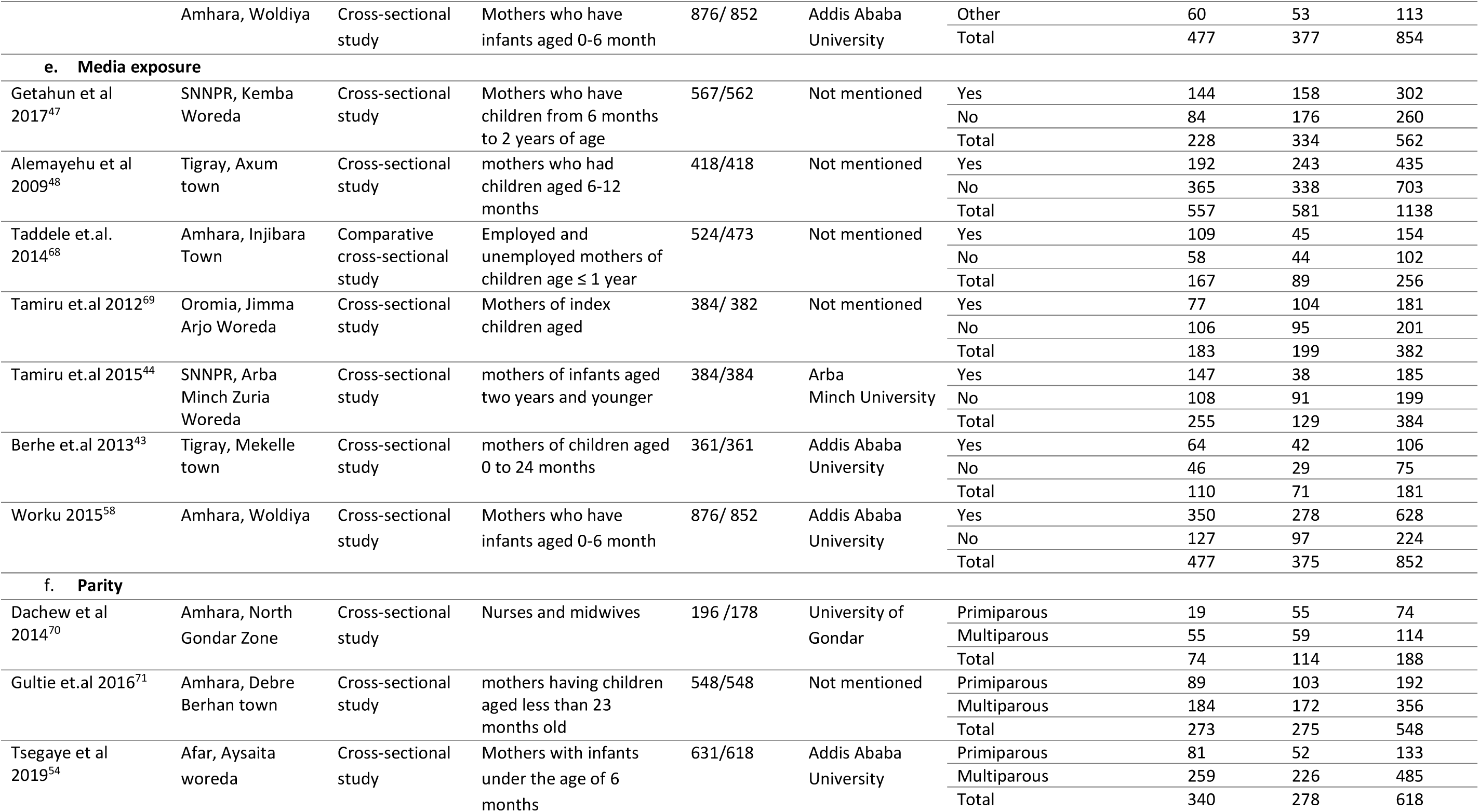
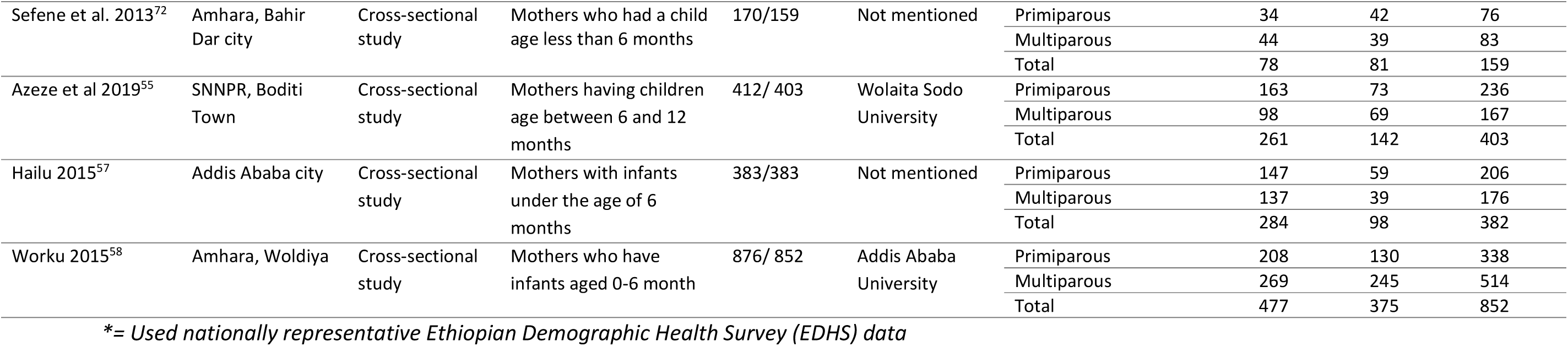
Summary of included studies

**Figure 1.**
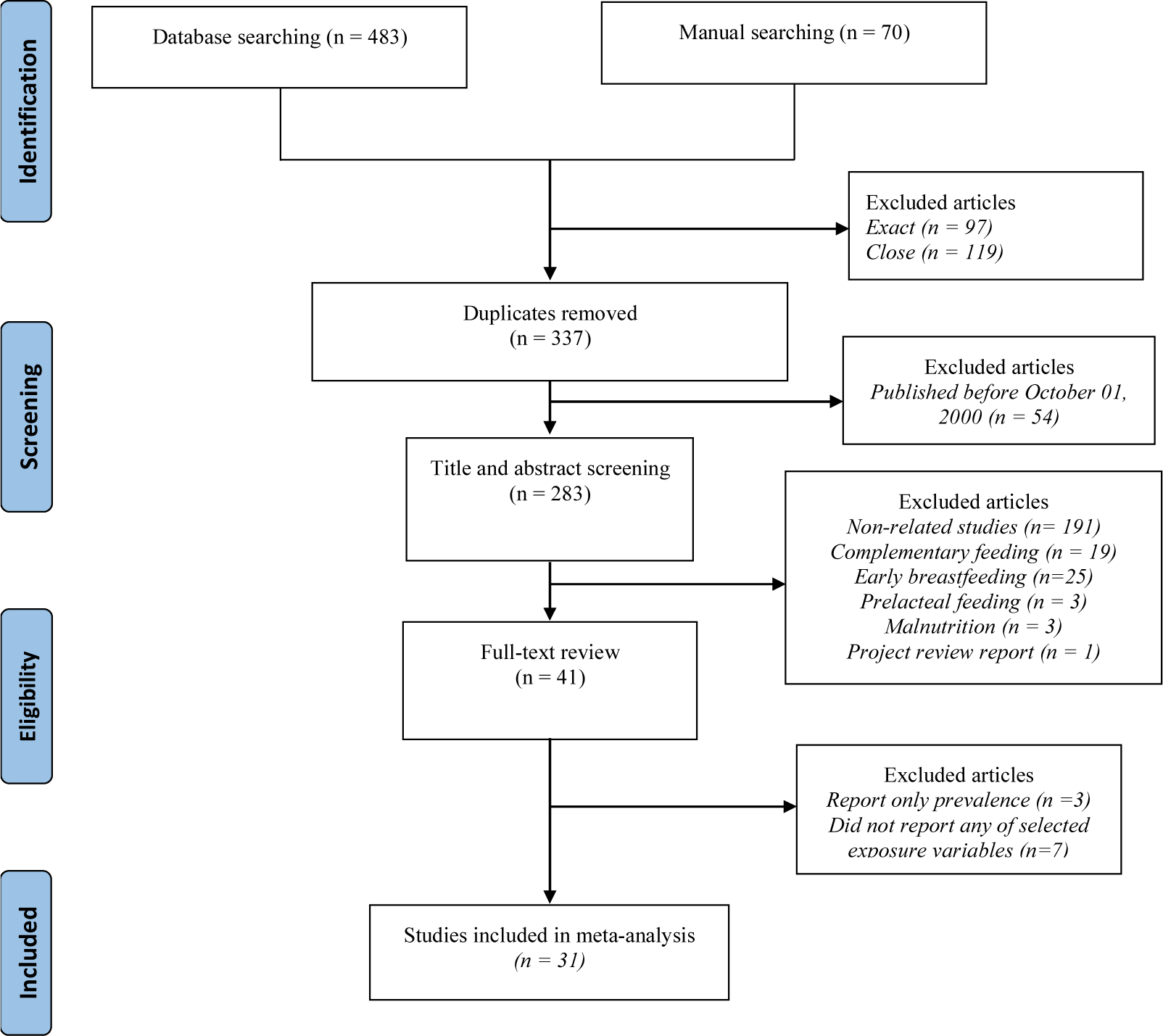
PRISMA flow diagram showing the schematics of literature screening and selection.

## Predicting factors

### Maternal educational status

Seventeen studies^42-58^ involving 14,691 mothers reported the association between maternal educational status and EBF (Table 1a). Seven studies were conducted in Amhara region, five in SNNPR, and four studies in Addis Ababa, Afar, and Tigray. One study used nationally representative data. The odds of EBF in mothers who attained primary education was 39% significantly higher than uneducated mothers (OR = 1.39; p = 0.03; 95% CI = 1.03 - 1.89; I^2^ = 86.11%) (Figure 1). This estimate was obtained after removing one outlier study.^58^ The meta-regression analysis result showed that region, sample size and publication year explain 24.54% of the between-study heterogeneity. These factors had also a moderation effect. The odds EBF among mothers with secondary education and above was 9% higher than uneducated mothers (OR = 1.09; p = 0.71; 95% CI = 0.69 - 1.72; I^2^ = 93.96%) (Supplementary Figure S3). The meta-regression analysis result showed that region, sample size and publication year explain 10.82% of the between-study heterogeneity, however, moderation effect was not observed (QM= 9.03, df = 7, p = 0.25). The odds of EBF in mothers with secondary education and above was 15% lower than mothers who attained primary education (OR = 0.85; p = 0.25; 95% CI = 0.64 - 1.12, I^2^ = 83.71% %) (Supplementary Figure S6). Based on the meta-regression analysis result, 67.13% % of the between-study heterogeneity accounted for the difference in region, study area, sample size and publication year. Region had moderation effect (QM= 26.47, df = 9, p = 0.002). In all three comparisons, there was no significant publication bias (Supplementary Figure S1, Supplementary Figure S4, Supplementary Figure S7) and the evidence on the effect of maternal educational status on EBF was mixed with steady (Supplementary Figure S2) and dramatic increment (Supplementary Figure S5 and Supplementary Figure S8) over time. Even though the association was not statistically significant at all levels, there was a clear pattern of a dose-response relationship between maternal educational status and EBF whereby the practice of EBF decreased when the educational status is increased.

**Figure 1:**
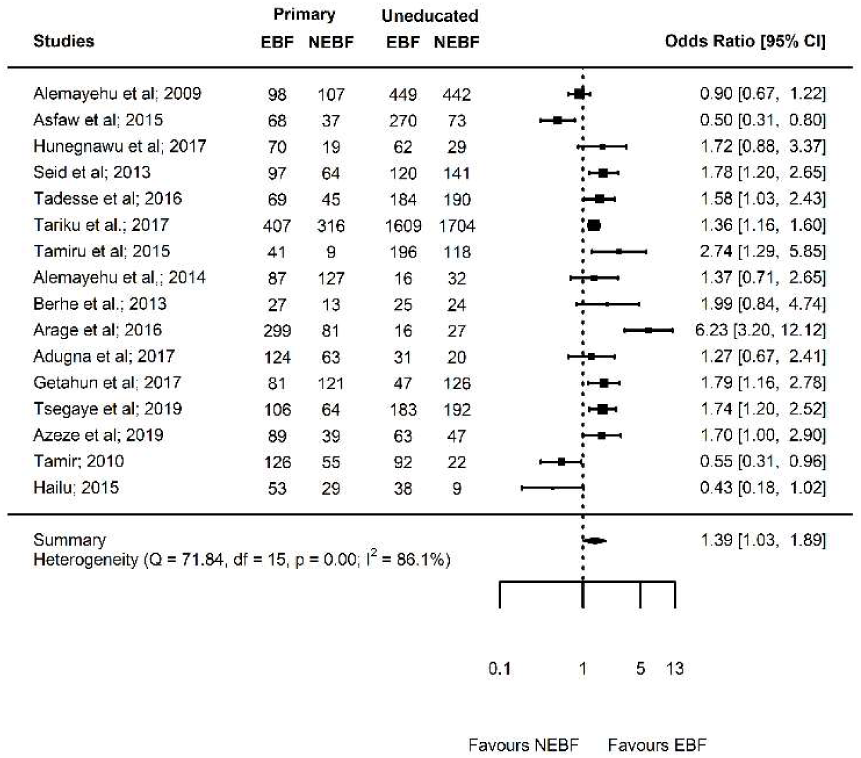
Forest plot showing the results of 17 studies examining the association between maternal educational status (primary education versus uneducated) and exclusive breastfeeding.

### Paternal educational status

Five studies^44,49-51,57^ with 2,698 mothers reported the association between paternal educational status and EBF (Table 1b). Three studies were conducted in Amhara region, and the other two studies conducted in Addis Ababa and SNNPR. Mothers who had spouse with primary education had 23% higher chance of EBF than mothers who had had uneducated spouse (OR = 1.23; p = 0.35; 95% CI = 0.80 - 1.90, I^2^ = 59.35%) (Figure 2). Likewise, mothers who had spouse with secondary education and above had 5% higher chance of EBF than mothers who had had uneducated spouse (OR = 1.05; p = 0.83; 95% CI = 0.67 - 1.66; I^2^ = 69.28%) (Supplementary Figure S11). Moreover, mothers who had spouse with secondary education and above had 9% higher chance of EBF than mothers who had had spouse with primary education (OR = 0.91; p = 0.46; 95% CI = 0.71 - 1.17, I^2^ = 0.00%) (Supplementary Figure S14). In all comparisons, there was no significant publication bias (Supplementary Figure S9, Supplementary Figure S12, Supplementary Figure S15). The evidence on the effect of paternal educational status on EBF was mixed with slight increment (Supplementary Figure S10 and Supplementary Figure S13) and decrement (Supplementary Figure S16) over time. Similar to maternal educational status, an inverse dose-response relationship was detected between paternal educational status and EBF although the association was not statistically significant at all levels.

**Figure 2:**
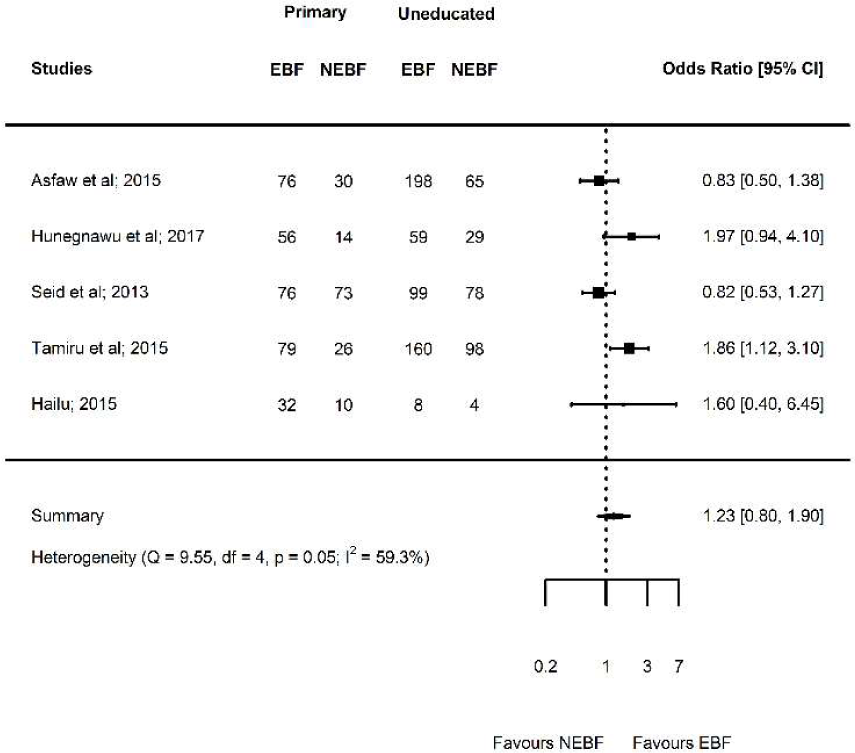
Forest plot showing the results of five studies examining the association between paternal educational status (primary versus uneducated) and exclusive breastfeeding.

### Household income

Thirteen studies^43,45,48-51,53,59-64^ with 11,966 mothers reported the association between socioeconomic status and EBF (Table 1c). Seven studies were conducted in Amhara region, two studies in Tigray region, and the other three studies conducted in Addis Ababa, Harari, and SNNPR. One study used nationally representative data. The odds of EBF among mothers who had medium socioeconomic status was 27% significantly higher than mothers who had low socioeconomic status (OR = 1.27; p = 0.02; 95% CI = 1.05 - 1.55; I^2^ = 60.9%) (Figure 3). The odds of EBF among mothers who had high socioeconomic status was 11% lower than mothers who had low socioeconomic status although not statistically significant (OR = 0.89; p = 0.47; 95% CI = 0.66 - 1.21; I^2^ = 83.11%) (Supplementary Figure S19). Based on the meta-regression analysis, 29.41% of the heterogeneity accounted for the variation in region, study area, study area and publication year, however, none of these factors had moderation effect (QM = 11.55, df = 9, p = 0.24). Moreover, the odds of EBF among mothers who had high socioeconomic status was 31% significantly lower than mothers who had medium socioeconomic status (OR = 0.69; p = 0.002; 95% CI = 0.54 - 0.87; I^2^ = 68.82%) (Supplementary Figure S22). There was no significant publication bias in all comparisons (Supplementary Figure S17, Supplementary Figure S20, Supplementary Figure S23). The evidence on the effect of socioeconomic status on EBF was not substantially changed over time (Supplementary Figure S18, Supplementary Figure S21, Supplementary Figure S24).

**Figure 3:**
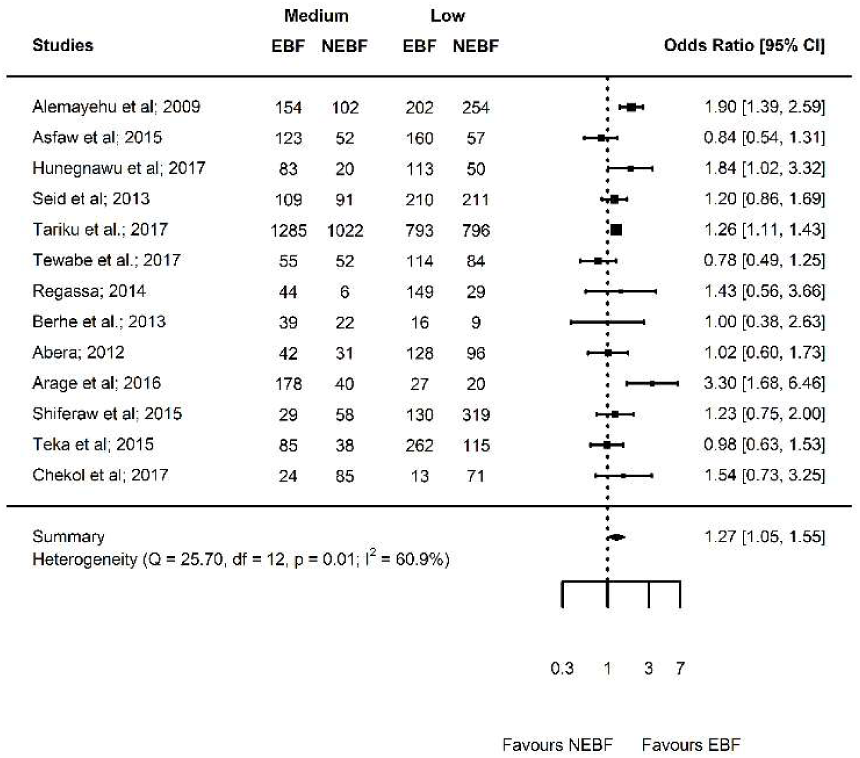
Forest plot showing the results of 13 studies examining the association between household income (medium versus low) and exclusive breastfeeding.

### Marital status

Sixteen studies^46,48,50,52-54,56-58,61-67^ including 15,689 mothers reported the association between marital status and EBF (Table 1d). Seven studies conducted in Amhara region, three studies in SNNPR and the five studies conducted in Addis Ababa, Afar, Harari, Oromia, and Tigray. One study used nationally representative data. One outlier study^48^ was excluded after sensitivity analysis. The odds of EBF among married mothers was 39% significantly higher than unmarried mothers (OR = 1.39; p = 0.02; 95% CI = 1.05 - 1.83; I^2^ = 76.96%) (Figure 4). There was no publication bias (Supplementary Figure S25). The evidence on the effect of marital status on EBF was slightly increased over time (Supplementary Figure S26).

**Figure 4:**
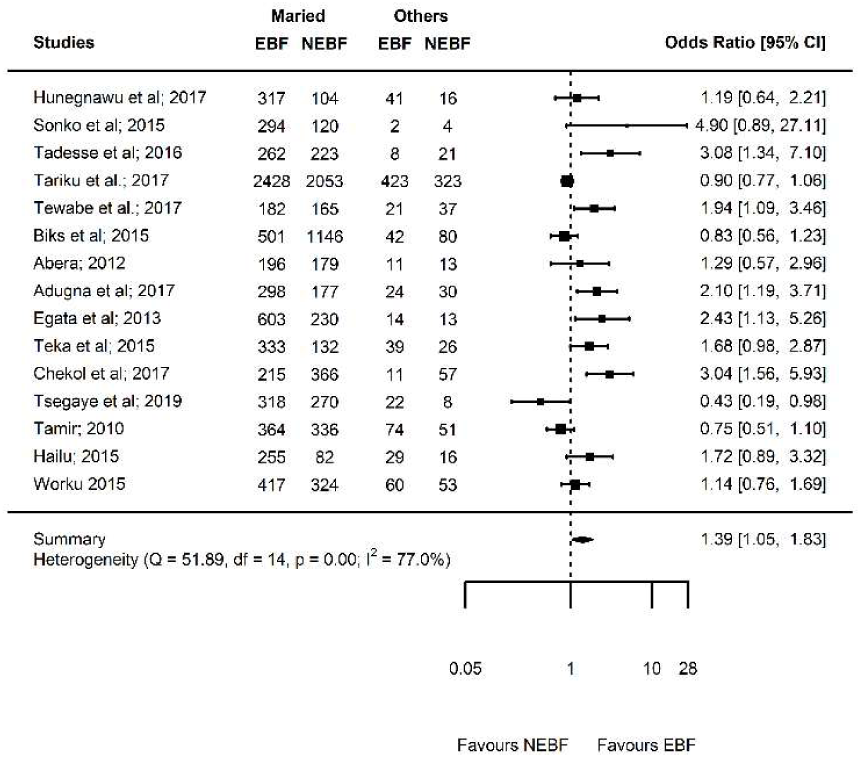
Forest plot showing the results of 16 studies examining the association between marital status (married versus others) and exclusive breastfeeding.

**Figure 5:**
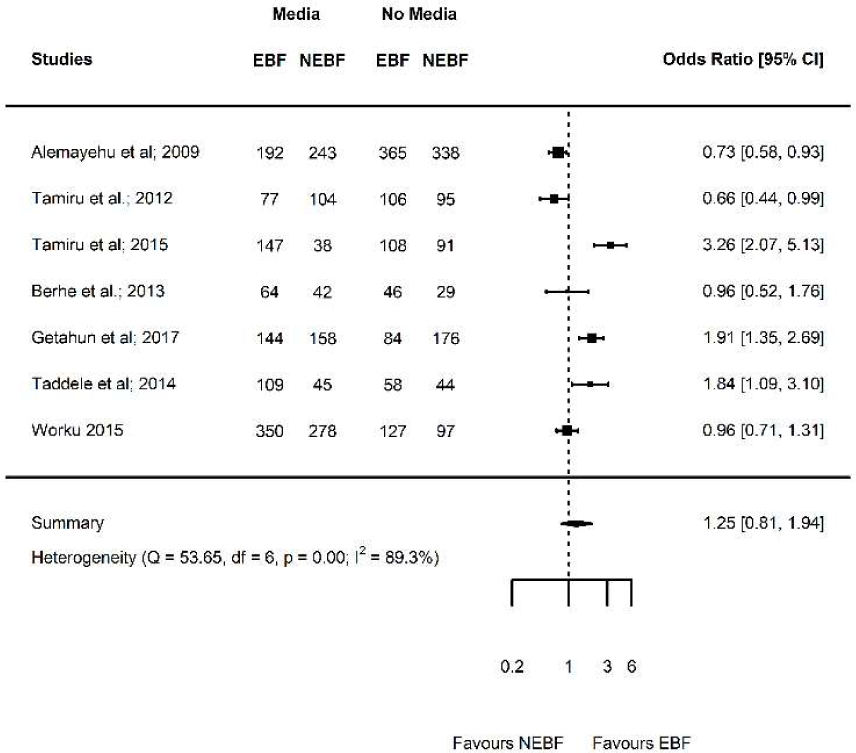
Forest plot showing the results of seven studies examining the association between media exposure (yes versus no) and exclusive breastfeeding.

### Media exposure

Seven studies^43,44,47,48,58,68,69^ in 4,231 mother reported the association between media exposure and EBF (Table 1e). Two studies conducted in Amhara region, two studies in SNNPR, and two other studies conducted in Oromia and Tigray region. One study used nationally representative data. The odds of EBF among mothers who had no media exposure was 25% higher than mothers who had no media exposure although the association was not statistically significant (OR = 1.25; p = 0.32; 95% CI = 0.81 - 1.94; I^2^ = 89.32%) (Figure 5). There was no significant publication bias (Supplementary Figure S27). The evidence on the effect of media exposure on EBF was steadily increased over time (Supplementary Figure S28). The meta-regression analysis result showed that 100% of the between-study heterogeneity accounted for the variation in region and publication year, and these factors had significant moderation effect (QM= 53.58, df = 5, p ≤ 0.01).

**Figure 5:**
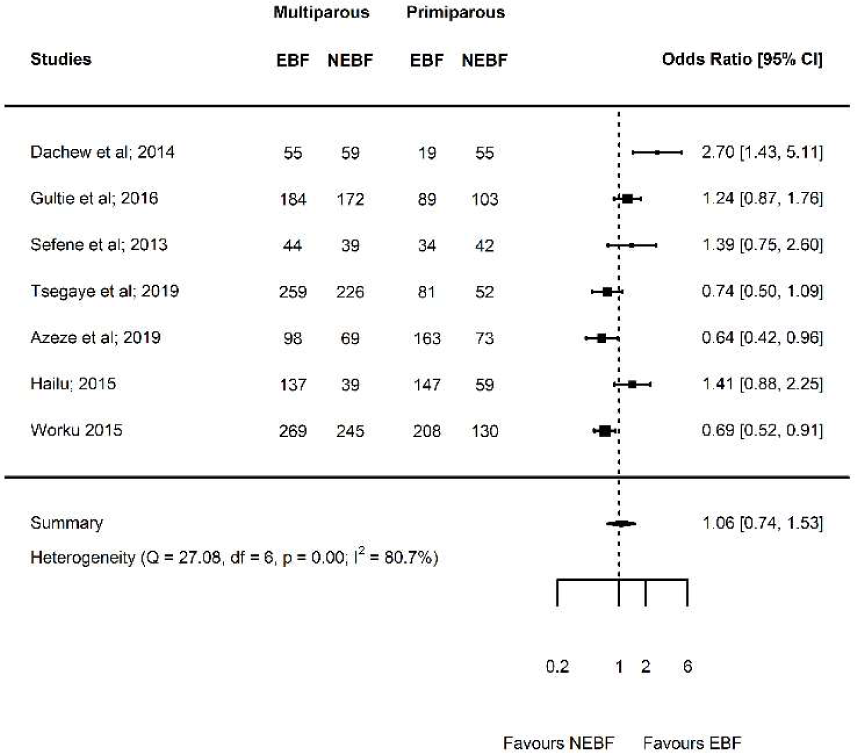
Forest plot showing the results of seven studies examining the association between parity (multiparous versus primiparous) and exclusive breastfeeding.

### Parity

Seven studies^54,55,57,58,70-72^ in 3,165 mothers reported the association between parity and EBF (Table 1f). Four studies conducted in Amhara region, and the other three studies conducted in Addis Ababa, Afar, and SNNPR region. The odds of EBF in multiparous mothers 6% higher than primiparous mothers although not statistically significant (OR = 1.06; p = 0.74; 95% CI = 0.74 - 1.53; I^2^ = 80.68%%) (Figure 6). There was a significant publication bias (Supplementary Figure S29). The evidence on the effect of media exposure on EBF was decreased over time (Supplementary Figure S30). The meta-regression analysis showed that 56.11% of the between-study heterogeneity accounted for the variation in the study area and publication year. Publication year had also moderation effect (QM= 6.94, df = 2, p = 0.03).

## Discussion

This meta-analysis investigated several factors that predicting EBF practice in Ethiopian mothers. The key findings were EBF significantly associated with maternal educational status, marital status and household income but not with parity, media exposure and paternal educational status. We further unraveled the dose-response relationship of educational status and household income and EBF although the association was not persistently significant at all levels.

Congruent with previous meta-analyses studies^73,74^, our meta-analysis showed that maternal primary education was beneficial for maintaining EBF until six months. This might be because mothers are more likely to be aware of the health benefits of breastfeeding if they are educated. Female literacy is essential to access health-related information, resist harmful traditional beliefs and practices, increase confidence and minimize early initiation of complementary feeding. On the other hand, in this meta-analysis, EBF among mothers having secondary education and above were not found to be significantly different from mothers with primary education as well as uneducated mothers. This finding suggests that the association between maternal education and EBF may not be linear. This implies only certain level of education may have a beneficial effect on EBF practice. This might be due to the fact that secondary and above education leading to better employment of the mother, engagement in intensive and time-consuming works which may buffer the relation between education and EBF. This hypothesis was supported by previous studies that found a negative correlation between maternal employment and EBF.^25,26,75^ Other studies also found mixed results on the association between maternal education and EBF. For example, in a review of studies conducted in Middle East countries, two studies found a significant positive association, other three studies revealed a significant negative association, and 10 studies could not confirm an association in either direction.^75^ In addition, a study based on the pooled Demographic and Health Survey data collected in nine Sub-Saharan Africa countries found that mothers with a secondary level of education or above were more likely to exclusively breastfeed compared with uneducated mothers.^26^

Another relevant finding was that currently married mothers had high chance of EBF compared with currently unmarried mothers. This might be due to several reasons. First, married mothers could have better incomes and educational levels^76^, as we showed certain level of education and household income advantage increased the likelihood of EBF. Second, previous study has shown that married mothers are more satisfied, committed to spousal relationships, get shared spousal support and reported less conflict. As a result, married mothers can have high emotional responsibility to keep the health of the newborn and more likely to engage in positive parenting behaviours.^77-79^ However, a systematic review of studies conducted in Brazil revealed only one out of six studies found a significant association between living with a partner and EBF.^80^

This meta-analysis also showed that mothers with a medium household income had higher chance of EBF than mothers with low household income. However, mothers with a high level of household income had lower chance of EBF than mothers with medium income and are not significantly different from mothers with low household income. Our result was in line with a study in five East and Southeast Asian Countries that showed a significant association between high household wealth index and low EBF practice.^73^ Even though a certain level of income is beneficial for EBF, it may decrease when the socioeconomic status is increased. This may be due to that mothers with high household income afford to and engage in formula milk, which lead to early initiation of complementary feeding. A previous meta-analysis depicted that low family income significantly contributes to the EBF interruption before six months.^74^ However, a large-scale study in nine Sub-Saharan Africa countries found no association between household wealth index and EBF.^26^

Furthermore, in this meta-analysis, parity, paternal education and media exposure were not found to be associated with EBF. This might be due to small number of studies included in the meta-analysis. Studies found a mixed finding regarding the association between parity and EBF. A meta-analysis of 22 epidemiological studies^74^ conducted in Brazil, and East and Southeast Asian Countries^73^ found that primiparity was associated with non-EBF. On the other hand, in a systematic review showed that only 6 out 19 studies found and a significant association EBF and multiparity.

This meta-analysis showed, for the first time, the pooled effect of several relevant predicting factors based on studies condcuted in the past 20 years. Even though residual heterogeneity is still evident, we managed this problem by conducting meta-regression analysis and by removing outlier studies using the Jackknife method. In addition, clinical heterogeneity was minimized by including only studies conducted on healthy mothers and newborns. Methodological heterogeneity was also minimized by including studies that reported EBF based on WHO definition, selected study subjects using a random sampling method, and similar study design and data collection methods. To minimize the possibility of missing relevant studies, we have used a combination of electronic databases search and manual search of cross-references, grey literature, and table of contents of relevant journals. Furthermore, this meta-analysis was conducted based on a published protocol to minimize methodological biases.

This meta-analysis has also limitations, which should be taken in to account during the translation of our results. All the studies included in this meta-analysis were cross-sectional study, which hinders inference on a cause-effect relationship. The risk of measurement error and recall bias should also be acknowledged. Interestingly, almost all included studies were conducted in mothers with a newborn less than 23 months. In relation to this, the maternal recall is found to be a valid and reliable estimate of breast feeding initiation and duration when the data is collected within three years of breast feeding history.^84^ Social desirability bias could also be evident given that self-reported breast feeding experience, educational status and income was used. In addition, the household income classifications are not standardized and did not account for indices such as inflation, which changes over time. Another problem was the confounding effect that can not be excluded given that the reported effect size in this meta-analysis was not adjusted to covariates, such as breast feeding counseling, cultural beliefs, partner support or maternal health status following childbirth. Studies for some variables were lacking in some regions, which may limit generalizability of our findings. Despite this fact, at least one study used nationally representative data per each variable. Similarly, only four studies investigated the association between paternal educational status and EBF, which may limits the statistical power of our meta-analysis. This meta-analysis covers studies condcuted only in Ethiopia; therefore, the results may not be extrapolated to other LMCs and developed countries. Moreover, statistical heterogeneity was observed in some of the analysis. The dose-response relationship of multiparity and EBF was not investigated due to the huge difference between included studies in the categorization of multiparity. A further limitation of this study was maternal and paternal education represents the formal education gained through schooling and it may not reflect the health literacy of the mothers and fathers. Finally, the results cannot be generalized to mothers and newborns with HIV/AIDS or other medical illness.

### Perspectives

Measuring breast feeding is challenging, and the use of standardized questions may be interpreted differently according to sociocultural contexts and differing probing techiniques by interviewers.^85^ In developing countries, breast feeding coverage is relatively good but we noted studies focused only on factors affecting breast feeding. To brideg this gap, future studies in Ethiopia should focus on investigating the effect of breast feeding on maternal and newborn health outcomes, such as cardiometabolic diseases, neuropsychological diseases, and psychiatric disorders. All included studies in the current review performed the traditional bivariate and multiple logistic regression. As a result, studies on associated factors should deeply consider the interconnection between predictors and apply detailed statistical analysis methods, such as mediation and moderation analysis. Moreover, future research should examine the possible causations between predicting factors and EBF practices in Ethiopia by implementing longitudinal research designs and large cohort-based studies, which can therefore, produce more accurate and insightful results. For example, one longitudinal study^24^ indicated the absence of significant association of several factors that consistently reported in cross-sectional studies. Overall, we observed a substantial inconsistency regarding predicting factors across nations; therefore, context-specific meta-analysis is further required to strengthen inferences. Given the scarcity of national, regional, or international meta-analysis that report the pooled effect of various relevant factors affecting EBF. future researchers in low- and middle-income countries should focus on meta-analysis instead of narrative review.

### Conclusions and recommendations

In this meta-analysis, we depicted the relevant effect of maternal education, income, and marital status on EBF. Therefore, multifaceted, effective, and evidence-based efforts are needed to increase national breastfeeding rates in Ethiopia. Health professionals should give special emphasis on uneducated or highly educated mothers with alternative supportive and educational interventions, such as prenatal education, counseling, and peer education.^81^ School education is also required for young girls to increase breast feeding awareness and to prepare for motherhood.^81^ Maternal education plays an important role in infant feeding behavioral change and to maintain the mother and newborn health. Even though there have been huge improvements in women education in Ethiopia, it has long been neglected and remains much lower than men. A systematic review of interventions designed to promote EBF in high-income countries found that the interventions using educational approaches were significantly associated with the increase in the duration of EBF.^89,90^ Another systematic review and meta-analysis showed that greatest improvements in early initiation of breastfeeding, exclusive breastfeeding and continued breastfeeding rates were seen when counseling or education was provided concurrently in home and community, health systems and community, health systems and home settings, respectively.^92^ Improving the economic power of women would also increase EBF practice to certain level. Health professionals should also give attention to unmarried mothers before, during and after birth. Workplace support for breastfeeding, minimizing cesarean section delivery, strengthening health-worker knowledge and skills at health facilities and Baby-Friendly Hospital Initiative, and strengthening the family- and community-level interventions are also relevant interventions.^25^ The training of health community agents and structural changes in health services might be relevant to increase EBF practice.^83^ Furthermore, improving the current maternity leave policy in Ethiopia that guarantees a pregnant female worker a paid maternity-leave of only three months of paid leave after delivery would be helpful to increase the rate of EBF. A woman’s ability to breastfeed is markedly reduced when she returns to work if breastfeeding breaks are not available, if quality infant care near her workplace is inaccessible or unaffordable, and if no facilities are available for pumping or storing milk.^87^ A study that explored the national policies of 182 Member States of the United Nations showed that the guarantee of paid breastfeeding breaks for at least 6 months was associated with an increase of 8.86 percentage points in the rate of EBF.^88^ Finally, access to breastfeeding support such as trained pediatricians, midwives, International Board-Certified Lactation Consultants, and peer counselors is considered important for supporting breastfeeding families, maternity, paternity, and parental leave, funding to support national and regional breastfeeding activities.^93^

## Data Availability

All data generated or analyzed in this study are included in the article and its supplementary files.

## Acknowledgment

Our special gratitude forwarded to Sjoukje van der Werf (University of Groningen, the Netherlands) for her support to develop the search strings.

## Contributors

TD conceived and designed the study. TD and BS developed the search strategy and screened studies. TD and SM extracted and analyzed the data and interpreted the results. TD and SM wrote the manuscript draft. All authors provided intellectual comments and revised the draft manuscript. All the authors read and approved the final manuscript.

